# Do health records data accurately identify repeat self-harm after emergency department visits?

**DOI:** 10.1101/2025.06.26.25330358

**Authors:** Gregory E Simon, Rebecca A Ziebell, Eric Johnson, Susan M Shortreed

## Abstract

**Objective:** Evaluate how often encounter diagnoses of self-harm soon after an emergency department visit for self-harm represent new self-harm events.

**Methods:** Electronic health records (EHR) and insurance claims data from a large integrated health system identified emergency department encounters for injury or poisoning coded as self-harm and then selected those with another encounter diagnosis of self-harm occurring within 91 days. Review of clinical text for these pairs of encounters examined whether the subsequent self-harm diagnosis represented a distinct new self-harm event vs. a repeat

**Results:** Of 121 pairs of encounters with relevant clinical text available for review, records indicated a distinct repeat self-harm event for 50 (41%, 95% CI 33%-49%). The proportion confirmed as distinct new events ranged from 3% (95% CI 0%-10%) for self-harm diagnoses the following day to 50% (95% CI 19%-81%) for diagnoses 2 to 7 days later to 100% (95% CI 92%-100%) for diagnoses 8 to 91 days later. The proportion confirmed as distinct, new events did not vary by healthcare setting of recording for the subsequent diagnosis (inpatient, emergency department, other outpatient) or similarity of injury or poisoning type between the two events.

**Conclusions:** Health systems, researchers, and public health agencies using insurance claims or EHR diagnoses to identify early recurrence of self-harm should be cautious regarding diagnoses that appear to represent early repetition of self-harm. Self-harm diagnoses the day after an emergency department self-harm visit rarely represent a distinct new event, while diagnoses recorded more than a week later usually do.

**HIGHLIGHTS:**

▸ Health records may over-estimate repeat self-harm after emergency department visit.
▸ Self-harm diagnoses the day after an ED visit rarely represent distinct new events.
▸ Self-harm diagnoses a week or more after an ED visit usually indicate new events.

## INTRODUCTION

Among people hospitalized for a self-harm event, approximately 1 in 60 will die by self-harm and 1 in 6 will experience another self-harm event over the following year (Carroll et al., 2014). Risk of a repeat event appears highest in the first days or weeks after self-harm (Fedyszyn et al., 2016). Identified risk factors for recurrence of self-harm include alcohol use and a history of additional prior self-harm events (Demesmaeker et al., 2021).

Bentley and colleagues, however, recently reported a potential pitfall in use of health records data to examine rates and predictors of repeated self-harm (Bentley et al., 2024). Among pairs of self-harm diagnoses in health records separated by less than 90 days, review of clinical notes found that over 80% of second diagnoses in each pair appeared to refer back to the prior event rather than indicating another distinct self-harm event. Diagnoses close together in time and those recorded in settings outside of the emergency department (ED) had an especially high rate of false identification of a new event. Those findings suggest that records data may substantially over-estimate risk of repeat self-harm events and may not accurately identify predictors or risk factors.

This report uses data from a large integrated health system to examine how often an emergency department visit for self-harm is followed by another diagnosis indicating an initial encounter for self-harm and how often those subsequent self-harm diagnoses accurately identify distinct new self-harm events.

## MATERIALS AND METHODS

Data were extracted from the research data warehouse of Kaiser Permanente Washington (KPWA), an integrated health system serving approximately 700,000 members in Washington state. KPWA members are enrolled via employer-sponsored insurance, individually purchased insurance (including subsidized insurance exchange plans for low-income members), Medicare, and Medicaid. Members are representative of the service area population in race, ethnicity, income, and educational attainment. The KPWA research data warehouse integrates data from electronic health records (EHR; for all services provided at KPWA facilities), insurance claims (for services provided outside of KPWA), and state mortality data. The KPWA Institutional Review Board approved a waiver of informed consent for use of deidentified medical records in this research.

The study sample included all ED and urgent care visits (index encounters) by KPWA members aged 11 or older between 10/1/2015 and 9/30/2017 with any diagnosis code(s) indicating an initial encounter for definite or possible self-harm, defined as ICD-10-CM codes for suicide attempt (T14. 91), injury with self-harm intent (X71-X83), injury with undetermined intent (Y21-Y33), poisoning (T36-T65) with modifier for self-harm or undetermined intent, and other poisonings or wound injuries accompanied by code for suicidal ideation (R45.851). ED self-harm visits leading directly to hospitalization were excluded.

For each index ED self-harm encounter, EHR and claims data identified all encounters starting 1 day later (the day after the index encounter) up to 91 days later, recorded in any setting (inpatient, ED/urgent care, or other outpatient) with any diagnosis indicating an initial encounter for definite or possible self-harm (as defined above). These pairs of self-harm diagnoses (index encounter and each subsequent self-harm diagnosis in the following 91 days) were then classified by number of days separating the two events, healthcare site of recording of the subsequent diagnosis (inpatient, emergency department, other outpatient), and similarity of diagnoses for the two events. Similarity of diagnoses was assessed at the level of the body region for injuries (e.g. S60-S69, injuries to the wrist, hand, and fingers) and category of substance for poisonings (e.g. T42, poisoning by antiepileptic, sedative-hypnotic, and antiparkinsonian drugs). For each pair of events, diagnoses were classified as identical (same categories of diagnoses recorded for index and subsequent event), partially overlapping (at least one diagnosis category the same and at least one different between the two events), and distinct (no shared or common diagnosis categories between the two events).

Pairs of index ED self-harm diagnoses and subsequent initial encounter self-harm diagnoses were selected for review of clinical text including a random sample of pairs 1 day apart, all pairs 2 to 7 days apart, and a random sample of pairs 8 to 91 days apart. A trained chart abstractor reviewed all available clinical notes on or around the date of the subsequent self-harm diagnosis and extracted any available text relevant to the occurrence or timing of a distinct subsequent self-harm event around that date as well as any text referring to a recent prior self-harm event. This extracted clinical text was then classified by the first author (GS, a practicing psychiatrist and user of this electronic health record system) who was blinded to the number of days between self-harm diagnoses pairs and the healthcare site of recording of the subsequent initial encounter diagnosis (all index diagnoses were recorded in the emergency department).

For each potential new event, relevant text was classified as either indicating a true distinct self-harm event or indicating a duplicate recording of the prior event.

Analyses examined the proportion of subsequent diagnoses classified as indicating true subsequent self-harm events, stratified by time between diagnoses, similarity of diagnoses, and healthcare site of recording of subsequent diagnosis. Confidence limits for proportions were estimated by exact binomial calculations, and test statistics comparing heterogeneity in proportions across strata were calculated using the Freeman-Halton extension of the Fisher exact probability test (Freeman & Halton, 1951).

## RESULTS

The criteria above identified 1544 ED index encounters by members aged 11 or older with an index visit with diagnoses of self-harm injury or poisoning. Of those index ED visits, 643 (41.6%) were followed by another encounter with a self-harm diagnosis within 91days, including 466 (30.2%) with self-harm diagnoses on the following calendar day, 72 (4.7%) with events 2 to 7 days later, and 105 (6.8%) with events 8 to 91 days later.

Of 254 pairs of self-harm diagnoses selected for review, no clinical text from any encounter was found on or near the subsequent event date for 67 (26%). This scenario likely reflects subsequent event encounters outside of KPWA for which an insurance claim was received but no clinical notes were shared. For an additional 66 (26%), some clinical text was available, but that text had no mention of *any* injury or poisoning. That scenario may reflect either a subsequent event encounter outside of KPWA for which no clinical notes were shared or erroneous recording of a self-harm injury or poisoning for an encounter not related to injury or poisoning. Relevant text was available for the remaining 121(48%) pairs of self-harm diagnoses.

Table 1 shows the proportion of subsequent events with relevant text classified as indicating a distinct self-harm event according to time between visits, healthcare setting of the subsequent self-harm diagnosis, and similarity of diagnoses between the two encounters. Probability that a second diagnosis represented an independent event varied significantly by time interval between events, ranging from 3% for diagnoses recorded 1 day later to 100% for diagnoses recorded 8 to 91 days later. Confirmation rates did not vary significantly by site of recording for the second diagnosis or similarity of diagnoses between events.

**Table 1.**
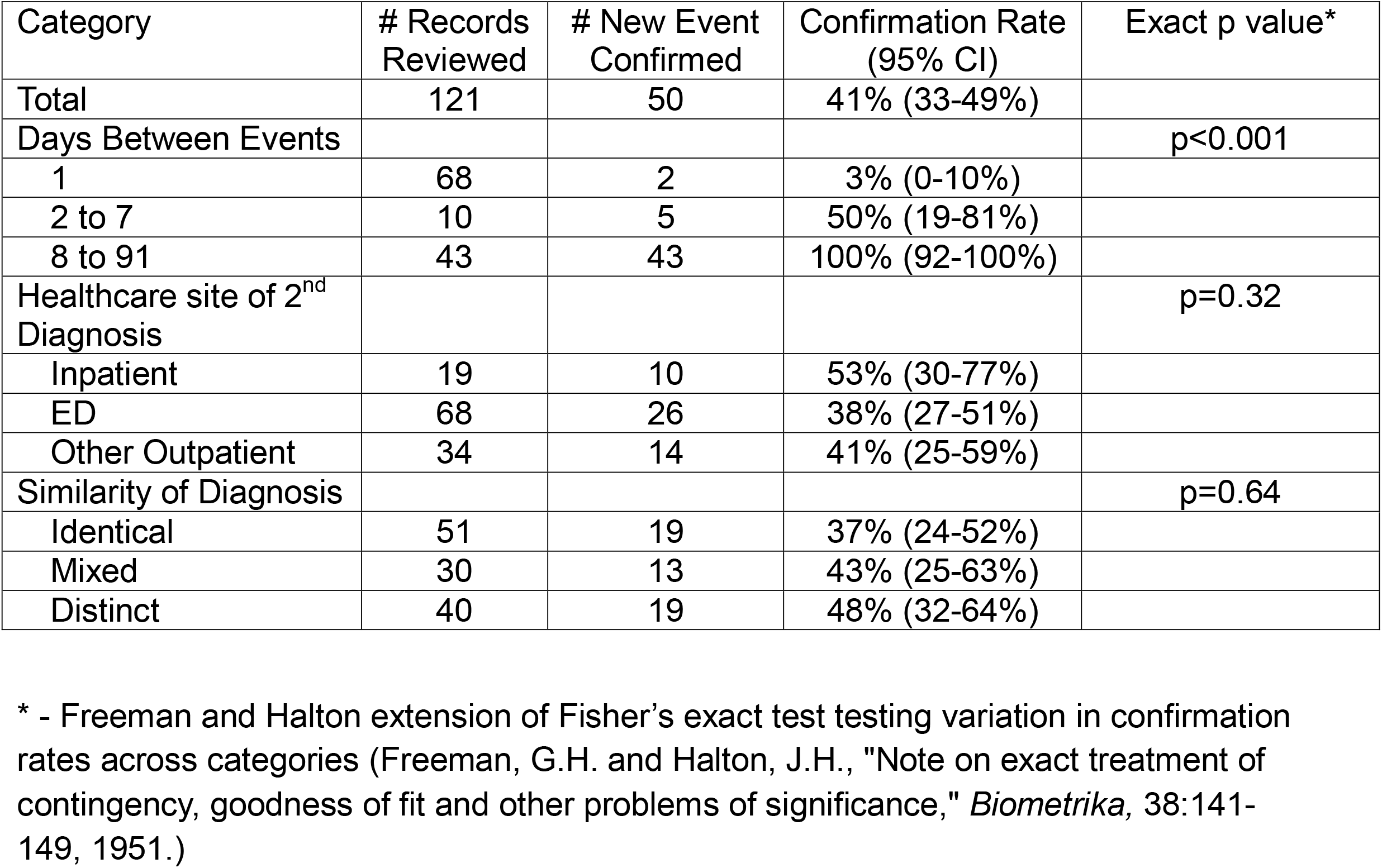
Proportion of self-harm diagnoses recoded after an emergency department visit for self-harm for which review of full-text records indicated a true distinct event.

## DISCUSSION

In this sample, encounter diagnoses from EHRs and insurance claims initially suggested a concerningly high rate of repeat self-harm (34.9% in the first 7 days after an emergency department visit for self-harm. Review of clinical notes indicated that most of those diagnoses recorded within 7 days actually referred to a prior event, suggesting a much lower true rate of new distinct self-harm events in the first week after an emergency department visit for self-harm.

Like Bentley and colleagues, we observe that the proportion of subsequent diagnoses accurately indicating a distinct new self-harm event varied markedly by time interval between the two diagnoses. Almost none (3%) of the subsequent diagnoses occurring the day after an ED visit for self-harm actually represented repeat self-harm, while all of those more than a week later accurately indicated a new self-harm event.

In contrast to the findings of Bentley and colleagues, we did not observe that the accuracy of coding for new self-harm events varied according to the healthcare site of the subsequent diagnosis. We also did not find that accuracy of coding varied by how similar or dissimilar pairs of diagnoses were to each other in terms of body location of injury and substance of poisoning.

While ICD-10-CM coding of injuries and poisonings requires specification of initial vs. subsequent encounter for any injury or poisoning event, we observed a high rate of subsequent encounters inaccurately coded as initial encounters. Clinicians may ignore that aspect of coding or may incorrectly use a “first encounter” code to indicate the first encounter with that specific clinician or facility rather than a first encounter for that injury or poisoning in any setting with any clinician.

Health systems, researchers, and public health agencies using insurance claims or EHR diagnoses to identify early recurrence of self-harm should be cautious regarding diagnoses that appear to represent early repetition of self-harm. Self-harm diagnoses the day after an emergency department self-harm visit rarely represent a distinct new event. In contrast, diagnoses of initial encounter for self-harm recorded more than a week after an emergency department visit for self-harm usually do represent new events.

## Data Availability

Data are not available, given risk that people with emergency room visits for self-harm could be identified.

## ACKNOWLEDGMENTS

Supported by NIMH Cooperative Agreement U19 MH121738

## DECLARATIONS OF INTERESTS

The authors have no relevant financial interests or other competing interests to declare.

## Notes

### Competing Interest Statement

The authors have declared no competing interest.

### Author Declarations

The Kaiser Permanente Interregional Institutional Review Board approved a waiver of informed consent for use of deidentified medical records in this research.

